# Classification of sinonasal pathology associated with dental pathology or dental treatment

**DOI:** 10.1101/2020.03.08.20032789

**Authors:** Beatriz Pardal-Peláez, José Luis Pardal-Refoyo, Javier Montero-Martín, José González-Serrano, Juan López-Quiles-Martínez

## Abstract

**Background:** As is known, the pathologies of the posterior teeth in the first and second quadrant and their treatments can be associated with pathology of the maxillary sinus. Both implant and pre-implant surgery have increased their incidence. It is necessary, therefore, to address sinonasal pathology (SN) related to dental pathology (DP) or dental treatments (DT) from an interdisciplinary point of view by establishing collaborative working groups between Dentistry (DEN) and Otolaryngology (ENT), as well as by developing registries and establishing coordinated diagnosis and treatment protocols of sinonasal pathology. The aim of this study is to design a useful classification of SN pathology associated with DP or DT to record information of DEN and ENT.

**Material and methods:** Bibliographic review and design of a classification system of SN, DP and DT pathology.

**Results:** Six categories are described in our system: 0- absence of SN pathology and DP; 1 and 4- patients with DP associated or associated with SN pathology, respectively; 2 and 5- patients with DT not associated or associated with SN pathology, respectively and 3- patients with SN pathology without DP. The classification has applications in diagnosis (association and possible causal relationship between the SN pathology and DP) and in the treatment of NS and DP simultaneously or sequentially.

**Conclusions:** The classification proposed integrates the presence or absence of DP or DT, and its causal association or the lack of it with SN pathology. Moreover, it facilitates the communication between DEN and ENT and eases the registration of information and the planning of dental, implant and pre-implant treatments.

## Introduction

The maxillary sinus (MS) is related to the antral teeth of the first and second quadrants (mainly premolars and molars and, eventually, canines). Thus, both dental pathology (DP) and invasive dental treatments (DT) can affect its structure (bone thickness, pneumatization, Schneider’s membrane, permeability of the osteomeatal complex) and its functions (ventilation, mucous secretion, mucociliary function). ^1-3^

The permeability of the osteomeatal complex is necessary for the physiology of maxillary sinus and may be affected by asymptomatic anatomical alterations or pathologies. ^4,5-7^

Alterations in mucociliary function may occur after any procedure in which there is manipulation of the membrane and may lead to pathology, mainly sinusitis, especially if it is associated with anatomical alterations of the maxillary sinus. This occurs in any preimplantation surgery that requires a good condition of the Schneider’s membrane. It is estimated that sinus membrane can be perforated in up to 52% of cases and, depending on its size, may or may not be repaired during surgery. ^8,9^

Invasive dental treatments can cause sinonasal pathology or other complications such as infections, oroantral fistula (from 5% to 25% after upper teeth extraction, intrasinusal foreign body, implant dislocation or periimplantitis. ^2,3,9^

As is known, 10% to 30% of maxillary sinusitis cases have dental origin –91% in molars–. In addition, maxillary sinusitis is the most frequent complication after invasive dental treatments –up to 27%–, although the diagnostic criteria are not always clear. ^9,10^ Approximately 4.17% (4/96) of cases of maxillary sinusitis are detected after implant placement and, out of these, 75% (n = 3) have a previous clinical history of sinonasal pathology. Therefore, it is advisable to collect the sinonasal semiology and make a preoperative otorhinolaryngological evaluation in patients who are going to receive dental implants in the antral areas. ^11^ Besides, up to 4.5% of patients undergoing sinus lift are likely to have an acute maxillary sinusitis, 1.3% of which will develop a chronic maxillary sinusitis. ^9^

Otolaryngological evaluation is necessary in maxillary implantation when symptoms of sinonasal pathology appear (endoscopic or radiologic), or in cases in which the sinus is going to be opened for bone augmentation (sinus lift). This is due to the fact that the state of the maxillary sinus will determine both implant and pre-implant surgery planning (including decisions about whether it will be performed or not), and will define not only the type of sinonasal treatment, but also the indication of the implant and the treatment sequence. ^9^

Consequently, implant surgery aimed at replacing antral teeth, (especially if the procedure needs sinus lift (pre-implant)), needs to be based on the evaluation and diagnosis of dental pathology or any complication sign of performed dental treatment, presence of sinonasal pathology and radiographic evaluation of maxillary sinus and dental by CBCT *(Cone Beam Computerized Tomography)*. ^12,13^ The CBCT allows the performance of sinonasal and dental exploration by evaluating the volume of the maxillary sinus, its pneumatization, ^7^ aeration, ^14^ bone thickness, ^15,16^ mucosa density, ^13^ the maxillary ostium and the osteomeatal complex permeability and the relation between the dental apex and the sinus floor. ^17^ Pneumatization of maxillary sinus can be altered by excess or, more commonly, by defect associated with various causes. ^7^ Between 61.8% and 66.9% of the radiological studies prior to implant surgery show sinonasal radiographic alterations that may have an impact on its results. ^4^

Table 1 summarizes some of the variables to be taken into account in dental and sinonasal exploration. ^6,7,13,14,16–18^

**Table 1.** Sinonasal and dental evaluation. Variables that should be registered.

|  |  |
| --- | --- |
| SINONASAL EVALUATION (6) | Nasal symptoms (rhinorrhea, nasal respiratory failure, epistaxis).<br>Sinusitis. Suspected acute, chronic or recurrent sinonasal pathology<br>History of previous nasal surgery<br>History of facial trauma<br>History of radiation in the head and neck<br>Findings in nasal endoscopy |
| DENTAL EVALUATION | Dental symptoms<br>Findings in dental exploration |
| SYSTEMIC PATHOLOGY | Diabetes, immunodeficiency, vasculitis, autoimmune pathology, smoking |
| RADIOGRAPHIC SINONASAL AND DENTAL EVALUATION (CBCT) (14) | Identifiable and permeable ostium<br>Signs of sinusitis<br>Signs of oroantral fistula<br>Pneumatization of the maxillary sinus(7) (hypoplasia or dilatation) |
| Tavelli et al. 2017 parameters (18) | RISK OF SCHNEIDER'S MEMBRANE PERFORATION<br>– MEMBRANE THICKNESS<br>– INTRASINUSAL TABICATIONS<br>– ANGLE BETWEEN BUCCAL AND PALATAL WALLS<br>– PRESENCE OF TEETH<br>– IMPLANTS OR TOOTH ROOTS NEAR SINUS |
|  | STATE OF THE BONE<br>– BUCCAL PLATE THICKNESS<br>– RESIDUAL HEIGHT OF ALVEOLAR EDGE<br>– WIDTH OF RESIDUAL ALVEOLAR RIDGE |
|  | OTHER PARAMETERS<br>– Sinus width<br>– Alveolar artery<br>– Visibility / oral opening |
| Mucosa of maxillary sinus<br>Tadinada et al. 2015 (13) | Class 1: Without pathology in maxillary sinus or thickness from 0-2 mm<br>Class 2: Thickness from 2-5 mm<br>Class 3: 6-9 mm with or without partial obliteration of maxillary sinus<br>Class 4: > 9 mm with partial or complete obliteration of maxillary sinus |
| Bone thickness<br>Juodzbalys et al. 2013 (16) | Type I: H >10mm<br>Type II: H (>4mm – ≤10mm)<br>Type III: H ≤4mm |
| Apex- sinus floor relation<br>Fry et al. 2016 (17) | Type 0 – Maxillary sinus floor over the molar<br>Type 1 – Tooth apex is in contact with maxillary sinus floor<br>Type 2 – Maxillary sinus floor is among the roots<br>Type 3 – The apex is inside the maxillary sinus |

Nowadays, the generalization of dental implant surgery techniques and reconstructive procedures for their positioning, together with the advances in imaging techniques (radiological and endoscopic) and in functional endoscopic sinus surgery (FESS), have increased the detection of sinonasal pathologies prior to implant surgery. They have also prompted the identification of sinonasal complications after implant surgery and the sinonasal interventions through FESS to correct anatomical anomalies, sinonasal pathologies or difficulties after invasive dental treatments that affect maxillary sinus. ^19^

These advances stress the need for collaboration between dentistry (DEN) and otoralyngology (ENT) in sinonasal pathology diagnosis, as well as throughout the planification process and in the performance of invasive dental treatments. Indeed, coordinated work would be beneficial during the whole process of implant surgery in the first and second quadrant, especially in pre-implant surgery, both in the preoperative phase, (including diagnosis and preventive treatment, when necessary, as well as guidance regarding the reversible or irreversible prognosis of sinonasal pathology that may condition the indication of the procedure), and during the operation in order to correct the pathologies, especially when it comes to re-establishing the physiological drainage of the maxillary sinus by medical or surgical treatment. Cooperation would also be of help in the postoperative period, particularly for the diagnosis and early treatment of possible complications of sinus lift procedures, maxillary sinusitis in particular. ^9,20^

Thus, it is crucial that the otoralyngologist (ENT) and the dentistist’s (DEN) objectives –namely restoration of the sinonasal function, and restoration of missing teeth and their function in chewing, swallowing, speeching and breathing, respectively– are unified by a simple classification system that relates both sinonasal (SN) and dental (DP) pathologies, or dental treatmets (DT) performed. This would allow an effective communication between professionals based on three aspects: otolaryngological diagnosis of anatomical or pathological sinonasal alterations (through endoscopy and radiological study), dental pathology diagnosis or dental treatment by the dentist as a possible cause of sinonasal pathology, and dental and sinonasal radiological diagnosis.

The aim of this paper is to propose a classification system that relates dental pathology or performed dental treatment with sinonasal pathology.

## Material and Methods

The project and design of the classification system began in December 2016 and was carried out by means of the following methodology:

1- Systematic review in databases –PubMed, Web of Science (WoS) and Cochrane Library– in search for articles that had been published between 1 January 2008 and 15 December 2019 was performed. Descriptors and strategies, which follow PRISMA guidelines, are summarized in Figure 1. ^21^

**Figure 1.**
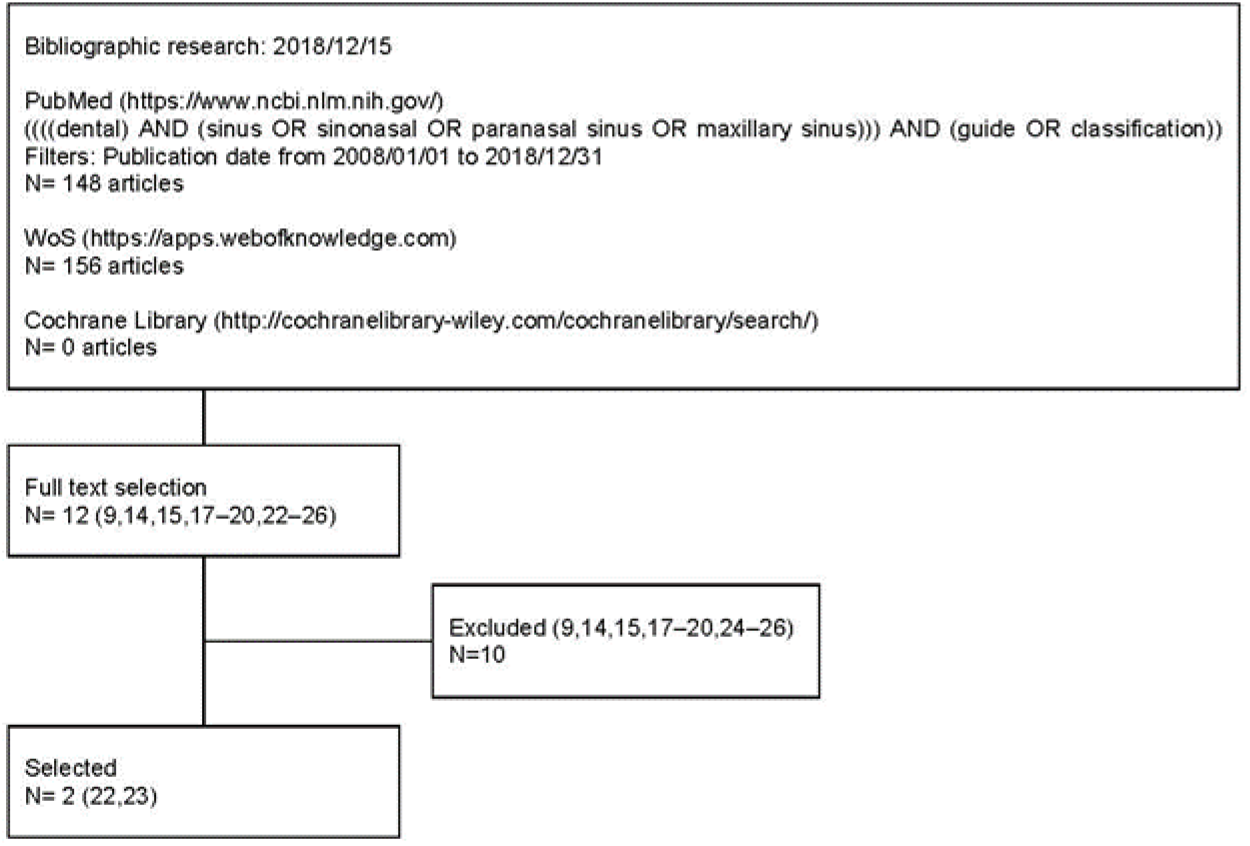
Summary of the descriptors and strategies in the bibliographic searches carried out following PRISMA guidelines.

As for inclusion criteria, articles in Spanish or English which analyzed sinonasal pathology and dental pathology or dental treatment performed in human adults were considered, together with those works including classification systems that relate sinonasal pathology and dental pathology or dental treatment.

Two of the authors performed independent searches and selected 12 full text articles that included classification systems. ^9,14,15,17–20,22–26^ Out of these, the two works which included a classification system of dental pathology or dental treatment related to sinonasal pathology were chosen. ^22,23^ The level of evidence was assessed by GRADE. ^27^

2- The decision was made that the system should fulfill the following purposes:

- Recording the results of the dental and sinonasal exploration.
- Recording the dental treatments performed and their possible complications.
- Allowing to relate both results of the records.
- Facilitating the recording of information within the clinical history of each patient.
- Facilitating the communication of information between dentist and otolaryngologist.
- Facilitating diagnostic and therapeutic decisions by dentist and otolaryngologist.
- Being adaptable to the classifications of diagnoses and treatments included in CIE9 and next.
- Being easy to remember.
- Admitting progressive revisions and updates.

3- The design of the classification system matrix that collects and relates sinonasal pathology and dental pathology or dental treatment performed was based on the reading of the selected literature (Table 2).

**Table 2.**
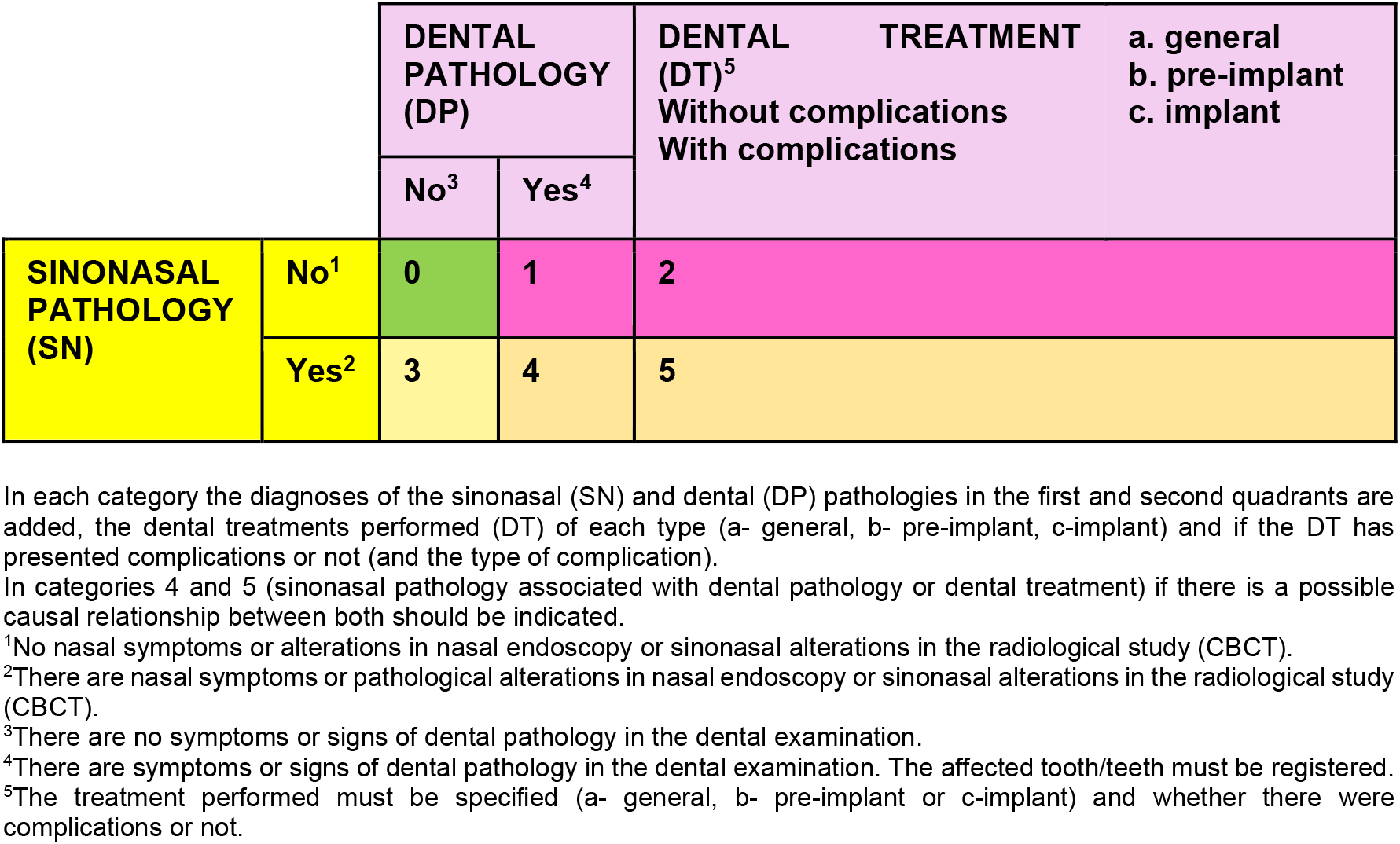
Matrix table for the classification design.

4- Review and modification of the successive versions of the classification.

The system versions were presented in three seminars on sinonasal pathology and implant surgery celebrated at the Faculty of Dentistry, Complutense University of Madrid, Spain (March 2017 and July 2018) and at the Faculty of Medicine and Dental Clinic, University of Salamanca, Spain (February 2018). An earlier version to the one proposed in this article was presented at the 69th Congress of the Spanish Society of Otolaryngology and Head and Neck Surgery and XVII Hispanic-Portuguese Congress of ENT in Madrid, Spain (October 2018).

A final review of the literature was made on December 15, 2019.

## Results

The classification system proposed in this paper is shown in Table 3.

**Table 3.**
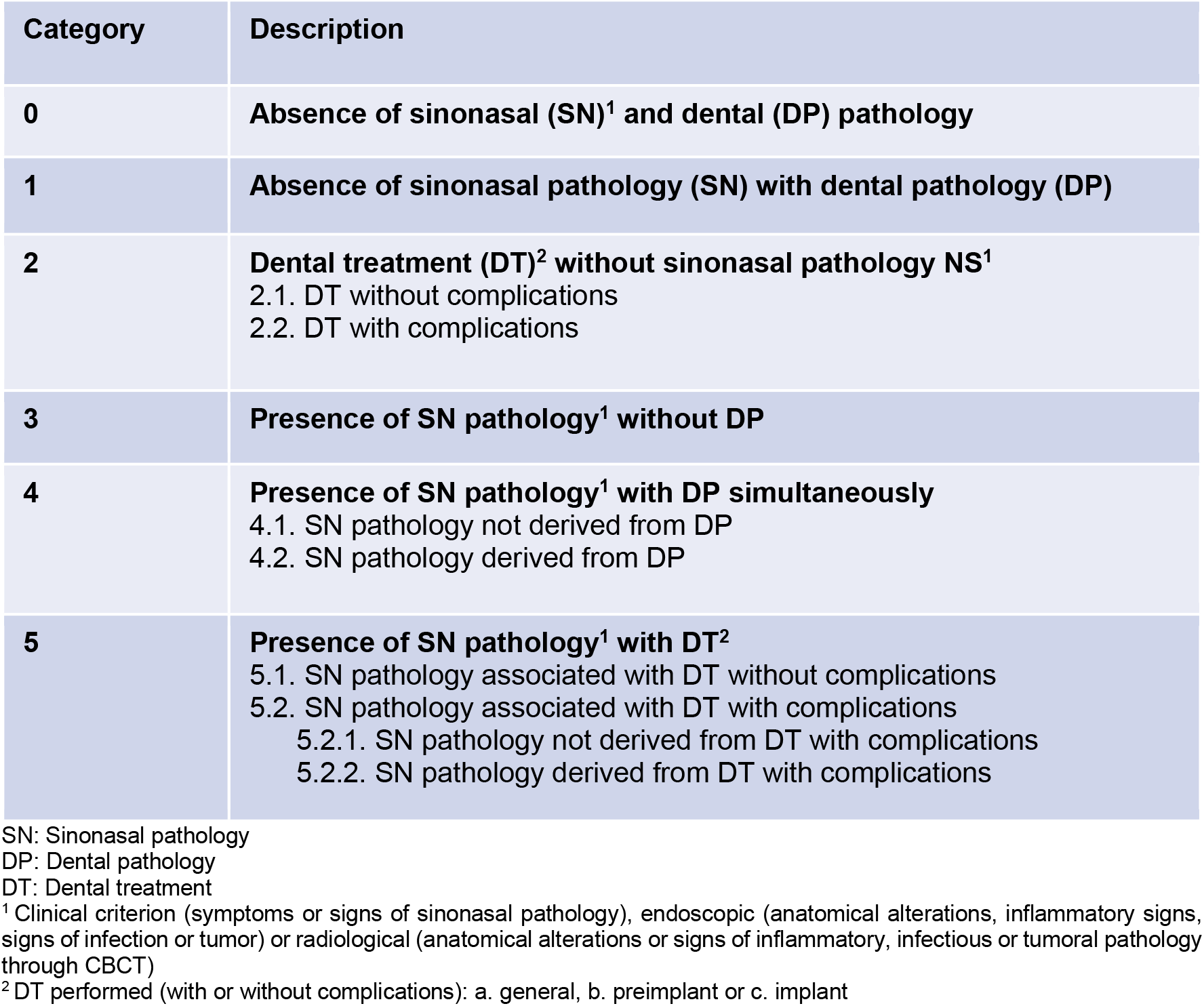
Classification of sinonasal pathology and its association with dental pathology or performed dental treatments.

| Category | Description |
| --- | --- |
| **0** | **Absence of sinonasal (SN)<sup>1</sup> and dental (DP) pathology** |
| **1** | **Absence of sinonasal pathology (SN) with dental pathology (DP)** |
| **2** | **Dental treatment (DT)<sup>2</sup> without sinonasal pathology NS<sup>1</sup>**<br>2.1. DT without complications<br>2.2. DT with complications |
| **3** | **Presence of SN pathology<sup>1</sup> without DP** |
| **4** | **Presence of SN pathology<sup>1</sup> with DP simultaneously**<br>4.1. SN pathology not derived from DP<br>4.2. SN pathology derived from DP |
| **5** | **Presence of SN pathology<sup>1</sup> with DT<sup>2</sup>**<br>5.1. SN pathology associated with DT without complications<br>5.2. SN pathology associated with DT with complications<br>5.2.1. SN pathology not derived from DT with complications<br>5.2.2. SN pathology derived from DT with complications |

SN: Sinonasal pathology
DP: Dental pathology
DT: Dental treatment
<sup>1</sup> Clinical criterion (symptoms or signs of sinonasal pathology), endoscopic (anatomical alterations, inflammatory signs, signs of infection or tumor) or radiological (anatomical alterations or signs of inflammatory, infectious or tumoral pathology through CBCT)
<sup>2</sup> DT performed (with or without complications): a. general, b. preimplant or c. implant

It includes six categories, numbered from 0 to 5, which aim at describing the different combinations possible depending on the existence or non-existence of sinonasal pathology and dental pathology or dental treatment.

The basic evaluation of the patient must include both otolaryngological (anamnesis and nasal endoscopy) and dental assessment (anamnesis and exploration) and radiological examination using cone beam computed tomography (CBCT).

Once results from basic assessment have been obtained, sinonasal pathology will not be considered in cases in which there are no nasal symptoms or alterations in nasal endoscopy or sinonasal alterations in the radiological study. It will be contemplated, though, if any of the former are detected. Similarly, dental pathology will be discarded unless the presence of any symptom or sign in dental or radiological exploration indicate otherwise.

As regards dental treatment, three possible groups are considered: a-general, b-pre-implant and c-implant. It is key to describe the type of dental treatment each patient should receive and whether there were complications during or after its completion.

Sinonasal alterations (anatomical, inflammatory, infectious or tumor), dental pathologies and radiological findings in CBCT are recorded for each patient.

## Discussion

As highlighted in this paper, bibliographical searches have revealed that only two studies have been developed which deal with a classification that relates the sinonasal pathology and the pathology and dental treatments ^22,23^; this points to a gap in the existing literature on the topic. The first article offers a classification from a retrospective design, and it analyses a mixed cohort of 257 patients with sinonasal pathology of dental origin or caused by dental treatments. However, it has no control groups, which limits the impact of the conclusions. ^22^ As for the second study, ^23^ the previous protocol was validated through a prospective study in a sample of 128 patients with sinonasal pathology caused by pathology or dental treatment that did not respond to adequate focal dental treatment. Therefore, the GRADE level of evidence was low or very low.

The classification described in Felisati *et al* ^22^ has diagnostic and therapeutic implications. It establishes three groups of maxillary sinus pathology caused by dental treatment from lower to higher risk of Schneider’s membrane injury –I-pre-implant surgery, II-implant surgery and III-general dental treatment– with seven classes that associate different complications derived from the treatments; these are infection, oroantral fistula, implant dislocation and peri-implant osteitis. This system relates the dental treatment (DT) (treatment group) with the complications (category) and is used by Kuan *et al*. ^10^

Based on this classification, we have developed a systematization protocol that relates sinonasal pathology and dental pathology or dental treatment without establishing a causal relationship, which allows us to classify all patients according to the existence or non-existence of dental pathology, sinonasal pathology and the antecedent of previous dental treatment, or the lack of it. Felisati *et al*’s ^22^ classification is implicitly included in category 5 of our classification.

As we have tried to show, the advantages of the proposed classification are the following:

– It includes all possible combinations when relating the existence or absence of MS pathology (sinusitis and other) and dental pathology, or the previous dental treatment (with or without complications) and the association of maxillary sinus pathology and dental pathology and dental treatment without presupposing its causal relationship. In each specific case, this potential relationship will be assessed.
– Category 0 is interesting in patients who require dental and otolaryngological assessment (for example, in cases of facial pain) but present no pathology.
– It is useful for the planning of treatments on sinonasal and dental pathologies simultaneously or sequentially. Categories 1 and 2 falls within the scope of dentistry, while ENT specialists would deal with category 4. Categories 4 and 5 would need unified and individualized dental and otolaryngological criteria for each patient, both in the diagnosis and in the treatment of each individual case (simultaneous or sequential).
– It is easy to remember and reproduce. It considers two sources of information (the existence or absence of sinonasal pathology and of previous dental pathology or dental treatment).
– It allows the improvement of the registration and communication between the dentist and the otolaryngologist.
– It helps the collaboration between the dentist and the otolaryngologist, both in the diagnosis and in the treatment in all stages of the implant surgery process: from the diagnosis and planning of dental treatment (preoperative stage), during dental treatment (operative stage), and until possible post-implant complications (postoperative stage). ^9^

## Conclusion

In conclusion, this paper has attempted to show how the proposed classification may allow us to improve the communication and registration of the pathologies of patients with sinonasal disease associated with pathology or previous dental treatment by involving professionals in dentistry and otorhinolaryngology. It can also aid in therapeutic decisions, while paving the way for subsequent prospective validation studies.

## Data Availability

All information is published in the article

